# Cost-effectiveness of tuberculosis infection screening at first reception into English prisons: a model-based analysis

**DOI:** 10.1101/2025.01.11.24319317

**Authors:** Nyashadzaishe Mafirakureva, Rachael Hunter, Claire F. Ferraro, Steve Willner, Thomas Finnie, Andrew Hayward, Andrew Lee, Anjana Roy, Chantal Edge, Peter J. Dodd

## Abstract

**Background:** The World Health Organization recommends systematic screening for tuberculosis in incarcerated populations, which are consistently at high risk of tuberculosis relative to the general population. In England, new receptions into prisons do not receive screening for tuberculosis infection, and evidence from economic evaluations is lacking.

**Methods:** We performed a cost-effectiveness analysis of introducing systematic screening for tuberculosis infection at first reception into English prisons from a health systems perspective. We used a tuberculosis transmission model calibrated to public data on prison stocks and flows. We developed decision tree models of prison-specific tuberculosis care pathways and their costs, informed by stakeholders and pilot studies. Sensitivity analyses included eliminating loss to follow-up (LTFU) in care cascades, zeroing extramural escort costs, and targeting screening to those born in countries with higher tuberculosis incidence (over 40 per 100,000 per year).

**Findings:** In our base case analysis, the intervention had an incremental cost-effectiveness ratio (ICER) of £78,000 per quality-adjusted life-year (QALY) gained. Reducing LTFU and avoiding prison escort costs would substantially improve cost-effectiveness, to ICERs of £70,000 and £54,000 per QALY gained, respectively. Targeting those born in higher incidence countries was predicted to be cost-saving.

**Interpretation:** Universal tuberculosis screening and preventive treatment for new receptions into English prisons is not cost-effective by the usual threshold of £30,000. However, targeting high-risk groups could be cost-saving. Tuberculosis interventions should explore ways to reduce LTFU and extramural healthcare in order to meet the needs of those incarcerated while minimizing costs.

**Funding:** UKHSA

## Introduction

Incarcerated populations are consistently at high risk of developing tuberculosis compared to the general population, with incidence rate ratios over 10 in all World Health Organization (WHO) regions.^1^ The global incidence of tuberculosis in incarcerated populations has been estimated at 125,000 per year,^2^ representing around 1% of the global total. In central and South America, rising incarceration rates mean that over 10% of notified tuberculosis is now among incarcerated people,^3^ and modelling suggests that changes in incarceration policy could contribute to renewed declines in tuberculosis for many countries in this region.^4^ While in the European region, tuberculosis notifications in incarcerated populations are declining, tuberculosis treatment outcomes remain substantially worse than in the general population,^5^ with the closed, crowded, communal nature of the setting highly conducive to the spread of respiratory disease.^6,7^ The resident population is also at higher risk of experiencing health inequalities, including less access to community healthcare, higher rates of tuberculosis risk factors such as rough sleeping and injecting drug use, and poorer health outcomes.^8,9^ Since 2021, the WHO has strongly recommended systematic screening for tuberculosis disease in incarcerated populations,^10^ but challenges to implementation remain,^11^ and recommendations and evidence on interventions for tuberculosis infection in incarcerated populations are lacking.

England is a WHO low tuberculosis incidence country, with a notification rate of 8.5 per 100,000 in 2023.^12^ There is however large variation by region and risk group, with 80% of notifications in those not born in the United Kingdom (UK), and 17% in those with social risk factors. In England in 2023, 4.2% of notified tuberculosis in people aged 15 years or over reported current or previous imprisonment.^12^ This estimate may well underestimate the contribution of prisons due to stigma in reporting imprisonment. Between 2017-2024, local Health Protection Teams in England and Wales have produced at least six tuberculosis prison outbreak reports, detailing high transmission rates and secondary cases, including to prison staff. Biobehavioural survey data from 2022-2024 across a variety of English prison types, and a previous a small-scale survey in a remand prison found tuberculosis infection rates of around 7%.^13^

Currently in England all new receptions into prisons are offered verbal symptom screening for tuberculosis disease; there is no routine assessment for tuberculosis infection. If individuals screen positive on the verbal screen for tuberculosis disease they will be isolated and referred for further evaluation including chest x-ray and molecular diagnostic testing of sputum. Challenges exist to this screening process, including symptoms going unrecognised or being discounted, for example due to long term smoking or drug withdrawal. In addition, operational pressures may limit capacity for isolation and influence decisions. Those requiring chest x-ray will usually be escorted by prison officers offsite to a community x-ray facility, which has associated National Health Service (NHS) costs, can be operationally challenging, and also offers a poor patient experience.^14,15^ Those who are found to have tuberculosis disease will be treated, and contact tracing undertaken to identify any associated cases, which may include screening contacts for tuberculosis infection.

While prisons are an important focus of national elimination strategies for tuberculosis in England and UK National Institute for Health and Care Excellence (NICE) recommends tuberculosis infection screening,^16^ there is no published evidence available on the cost effectiveness of screening and treating tuberculosis infection amongst people in English prisons. It is hypothesised screening in this population could identify those who would benefit from treatment, reducing the risk of developing tuberculosis disease and subsequent transmission and outbreaks, as well as reducing poor patient outcomes. This in turn would contribute more broadly to the national tuberculosis elimination agenda.

We therefore undertook a model-based cost-effectiveness analysis of tuberculosis infection screening for those entering pre-trial custody, compared to a standard of care where no screening takes place.

## Methods

Our modelling framework combined a model of the flow of incarcerated people through the English prison system, a model of tuberculosis transmission and illness, and models of patient care pathways to calculate changes in resource use, costs, and health outcomes under the intervention.

### Model of flow through the English prison system

To represent the distribution of durations spent detained in the English prison system system, we developed a stock and flow ordinary differential equation (ODE) model of the incarcerated population, and those released (Figure 1A). Under an equilibrium assumption, we fitted this model in a Bayesian paradigm to a number of targets identified from public data, namely: the total population; the fraction detained in open prisons; the total rates of release and sentencing; and the mean detention duration at release. The likelihood for each target was taken as normal with a standard deviation of 5% of the target value, which was used as the mean. Inference was performed using Markov chain Monte Carlo (MCMC) using Stan, and 4000 joint samples of the flow parameters were retained for use in analysis. For details of priors and parametrization, see Appendix.

**Figure 1:**
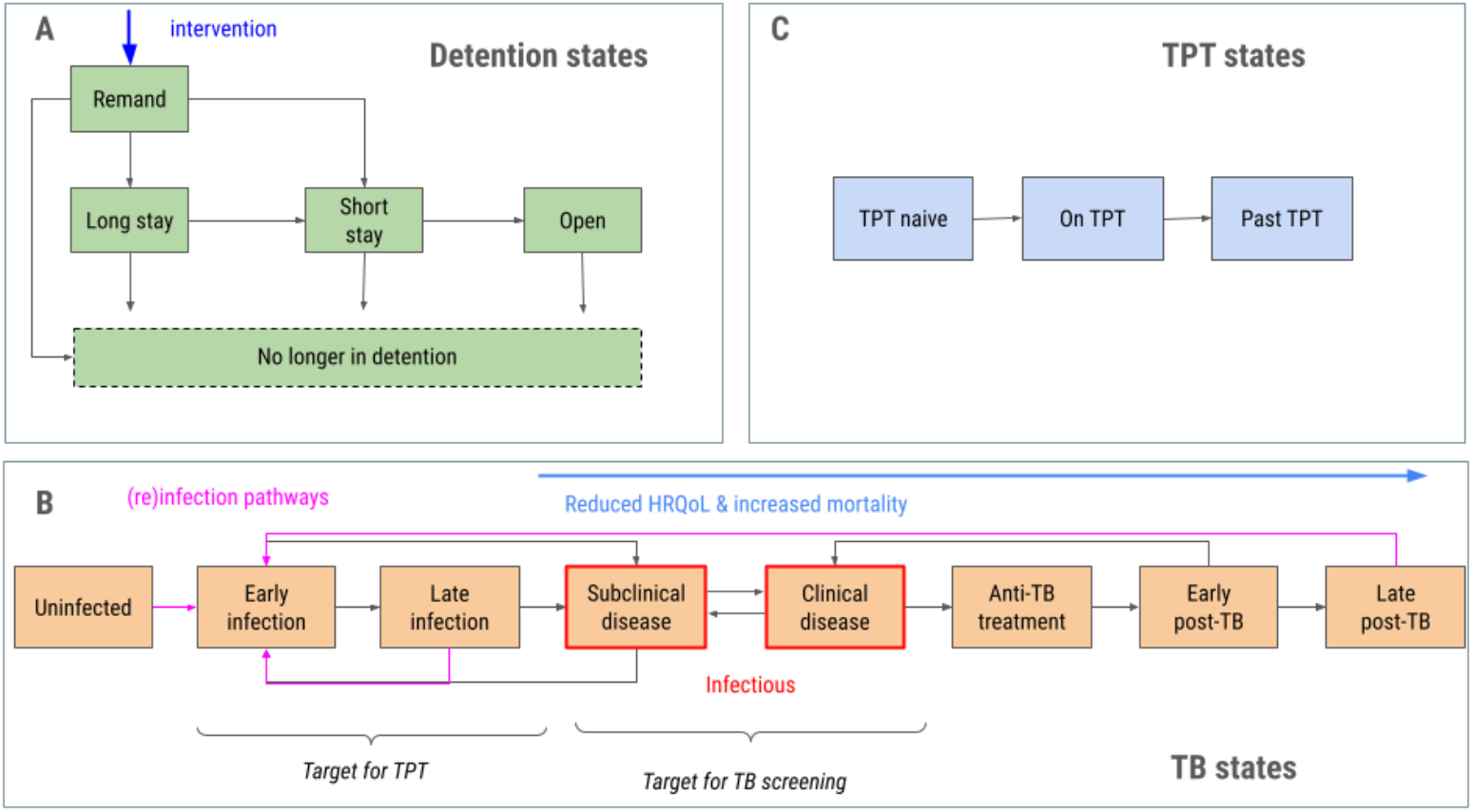
Transmission model structure. A: Detention states and flows between them. The intervention is situated at the flow into remand (blue). B: The tuberculosis (TB) states and transitions, with infectious states in red. States under the blue arrow experience worse health-related quality of life (HRQoL) and increased risk of tuberculosis or death. C: States representing tuberculosis preventive therapy (TPT). The full compartmental model is the product of these three domains, comprising 5 x 8 x 3 = 120 ordinary differential equations.

### Tuberculosis transmission model

To represent the natural history and transmission of tuberculosis, we extended our ODE model of the incarcerated population to include standard tuberculosis states, including subclinical and clinical tuberculosis disease, but also tracking those with previous tuberculosis disease as experiencing worse health-related quality of life (HRQoL) and higher risks of tuberculosis disease or death (Figure 1B). We further stratified the ODEs by tuberculosis preventive therapy (TPT) status (naive, current, previous; Figure 1C), for a system of 5 x 8 x 3 = 120 ODEs. Tuberculosis transmission within the prison system is assumed to be driven by untreated tuberculosis disease among those imprisoned, with random mixing. Community transmission once released is represented as a single generation of transmission and disease. Sensitivity analyses consider a fixed (static) force-of-infection in incarcerated populations and neglecting community transmission. For full details, see Appendix.

### Decision tree models of care pathways and intervention

To estimate the economic costs and outcomes of care, we developed decision tree models based on UK clinical and public health guidelines and discussions with practitioners. Three separate but linked pathways were represented: 1) screening for tuberculosis on first reception at prison; 2) tuberculosis infection screening and management pathway; and, 3) management of imprisoned people who develop tuberculosis symptoms.

The initial tuberculosis risk assessment should be done within 48 hours of arrival by verbal symptom screening and can include a chest x-ray where facilities exist, although x-ray rarely happens in practice. People who screen positive are evaluated for tuberculosis disease by physicians, typically involving clinical assessment, laboratory investigations and possibility for referral to the local NHS tuberculosis service. Individuals diagnosed with tuberculosis disease should start anti-tuberculosis therapy (ATT); those for whom tuberculosis disease has been excluded enter the second pathway, which may result in their initiating tuberculosis preventive therapy (TPT) if diagnosed with tuberculosis infection. The final pathway applies to those who develop tuberculosis symptoms while imprisoned, and may involve referral to local NHS tuberculosis services and may trigger contact tracing if tuberculosis disease is diagnosed. See Appendix for details.

Standard of care is assumed to involve low screening on imprisonment and is compared with complete coverage of screening (Figure 1A). Decision trees were operationalized in R, and for each input parameter set the fraction and cost of a cohort entering the pathway that ended in ATT, TPT, or no treatment were outputted, stratified by tuberculosis status (disease, infection but not disease, neither). These outputs were used to represent care pathway outcomes in the transmission model.

### Economic and analytical approach

The cost analysis primarily used a standard health system perspective following the NICE Technology Assessment guidelines,^17^ but identified and included prison service costs relevant to prison-based healthcare interventions. Resources required for activities along the care pathways were identified and the quantities of the resources estimated based on guidelines or literature. Resource use was valued by multiplying the resource use and relevant unit costs in 2021 Great British Pounds (£). Unit costs were obtained from relevant sources such as NHS reference costs, Personal Social Services Research Unit costs, prison-based pilot projects and literature. See Appendix for details.

We quantified health benefits as quality-adjusted life-years (QALYs) that summed life-years lost due to deaths caused by tuberculosis and reductions in HRQoL, during untreated tuberculosis disease and among tuberculosis survivors. We assumed a life-expectancy of 40 years.

Both costs and benefits were accrued over a 70 year time horizon after introducing the intervention with 3.5% discounting applied to both in base case. A cost-effectiveness threshold of £30,000 per QALY gained was used to demark cost-effectiveness and in calculating net benefit.

All model parameters were treated as uncertain within a probabilistic sensitivity analysis (PSA) framework. A sample with replacement of 10,000 parameters from the MCMC analysis of detention flows was merged with samples of the same size from decision tree outputs, and a sample of tuberculosis natural history and epidemiological parameters. These distributions are specified in the Appendix. For each parameter set, the transmission model was run initially for 50 years to avoid transients, and then costs and benefits captured under the intervention and standard of care. As analysis variants, we restricted our PSA samples to inputs that resulted in per capita tuberculosis notification rates among imprisoned people of between 30 and 100 per 100,000 person years.

We present results on the intervention TPT and ATT treatment cascades, and the proportion and average cost of TPT, ATT, or no treatment under intervention and standard of care for those with tuberculosis disease, infection, or neither. We also present total and incremental ATT and TPT courses, associated costs, and tuberculosis incidence, mortality and quality of life loss. Finally we computed net benefit and incremental cost-effectiveness ratios (ICERs).

### Sensitivity analyses and targeted screening

To understand the impact of particular assumptions, we ran sensitivity analyses in which we set to zero: cascade loss to follow-up (LTFU) for prison GP assessments, NHS attendance, treatment initiation, and treatment completion; cascade costs associated with escorts out of prison; community transmission; all transmission (fully static analysis); all transmission and all post-tuberculosis effects.

To consider the effect of targeting tuberculosis screening we modelled strategies targeting only higher risk groups among new receptions to prison. We parameterized the performance of two specific exemplar strategies highlighted based on biobehavioral survey data among imprisoned people, namely screening only those: 1) born in countries with estimated tuberculosis incidence >= 40/100,000 person-years; or 2) additionally, those with a history of homelessness, injecting drug-use, or problematic alcohol use. We modelled targeting as a zero-cost pre-screen step and present net benefits across a range of sensitivities and specificities (see Appendix) using a threshold of £30,000/QALY gained.

## Results

Using data from pilot studies on enrollment and retention through the ATT and TPT cascades in our model, suggested that even with full coverage of screening, 31% of those eligible for ATT and 24% of those eligible for TPT would make it through to treatment completion (Figure 2). Under our sensitivity analysis assuming no LTFU, we found corresponding treatment completion rates of 69% and 44%, respectively.

**Figure 2:**
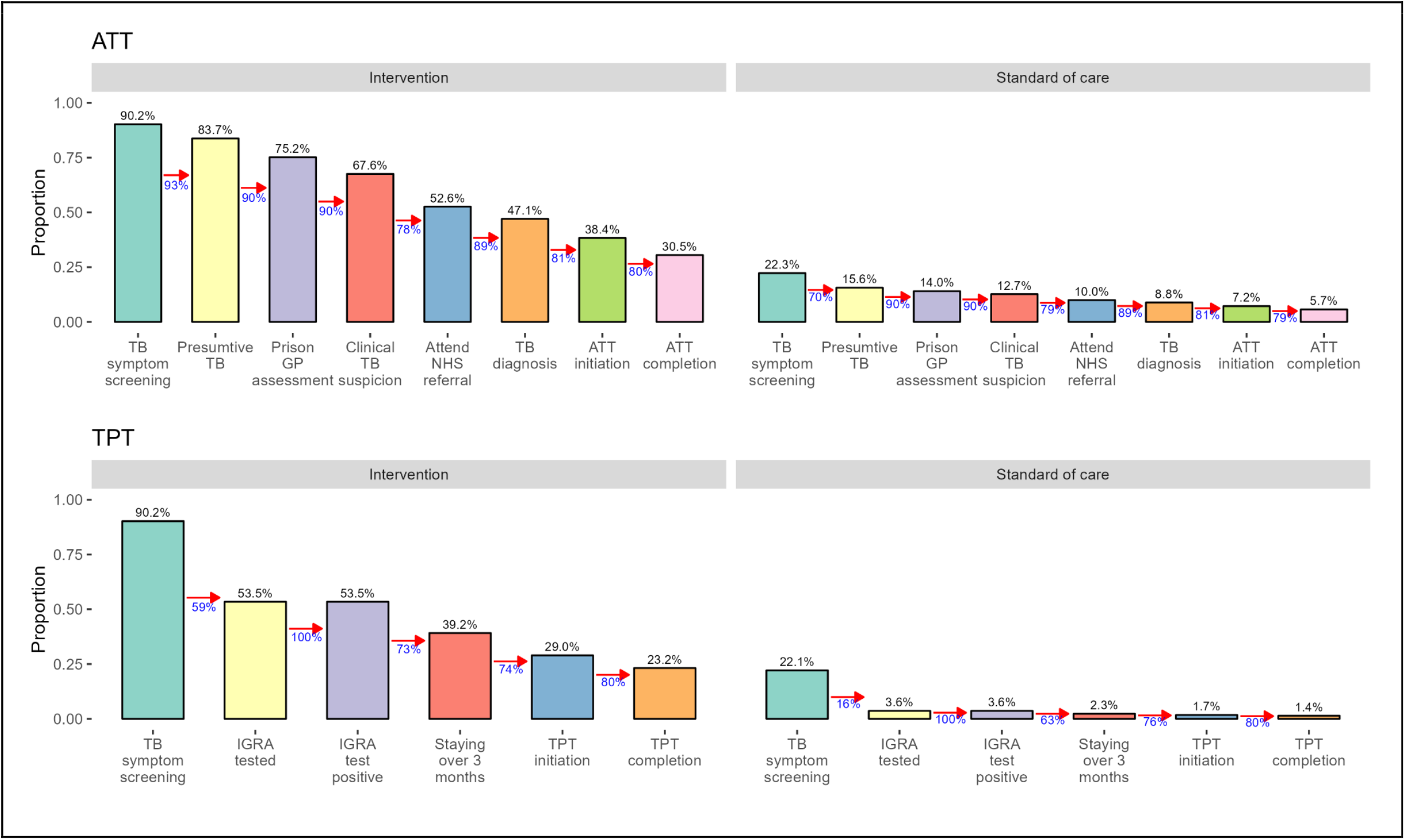
Intervention cascades. ATT = anti-tuberculosis treatment (for disease); TPT = tuberculosis preventive treatment; TB = tuberculosis; GP = general practitioner; NHS = National Health Service; IGRA = interferon gamma release assay (for tuberculosis infection)

The outcomes of these cascades are reflected in Table 2, which was used to represent the interventions in the transmission model. Table 2 also includes the probabilities of inappropriate treatments, as well as the unit costs for each tuberculosis state/treatment pair. Inappropriate treatment is rare, but often results in substantially larger unit costs due to typically longer routes through diagnosis to treatment.

**Table 1:** Screening pathway results. The cohort proportion and average cost by outcome and tuberculosis state at entry. Parentheses denote 95% quantiles around mean. TPT = tuberculosis preventive therapy; ATT = antituberculosis therapy; TB = tuberculosis. * excluding those on ATT or TPT ** excluding TB disease

| Arm | Quantity | Outcome\State* | TB disease | TB infection** | TB uninfected |
| --- | --- | --- | --- | --- | --- |
| Standard of care | Probability | ATT | 7.2% (1.8% to 16%) | 0.4% (0.1% to 0.9%) | 0.3% (0.1% to 0.7%) |
|  |  | TPT | 0% (0% to 0%) | 1.7% (0.1% to 6.2%) | 0% (0% to 0%) |
|  |  | neither | 92.8% (84% to 98.2%) | 97.9% (93.1% to 99.9%) | 99.7% (99.3% to 99.9%) |
|  | Unit cost | ATT | £28,305 (£23,558 to £34,009) | £45,092 (£35,291 to £60,447) | £44,775 (£34,830 to £61,738) |
|  |  | TPT | - | £9,199 (£4,691 to £21,773) | - |
|  |  | neither | £487 (£113 to £1,157) | £96 (£16 to £249) | £70 (£16 to £158) |
| Intervention | Probability | ATT | 38.4% (19.7% to 55.7%) | 1.2% (0.5% to 2%) | 0.8% (0.3% to 1.4%) |
|  |  | TPT | 0% (0% to 0%) | 29% (13.9% to 44.4%) | 0% (0% to 0%) |
|  |  | neither | 61.6% (44.3% to 80.3%) | 69.8% (53.9% to 85.3%) | 99.2% (98.6% to 99.7%) |
|  | Unit cost | ATT | £28,257 (£23,585 to £33,834) | £59,905 (£43,647 to £92,133) | £53,402 (£38,037 to £84,837) |
|  |  | TPT | - | £3,855 (£3,049 to £5,083) | - |
|  |  | neither | £3,953 (£1,755 to £7,649) | £794 (£485 to £1,191) | £240 (£188 to £296) |

**Table 2:**
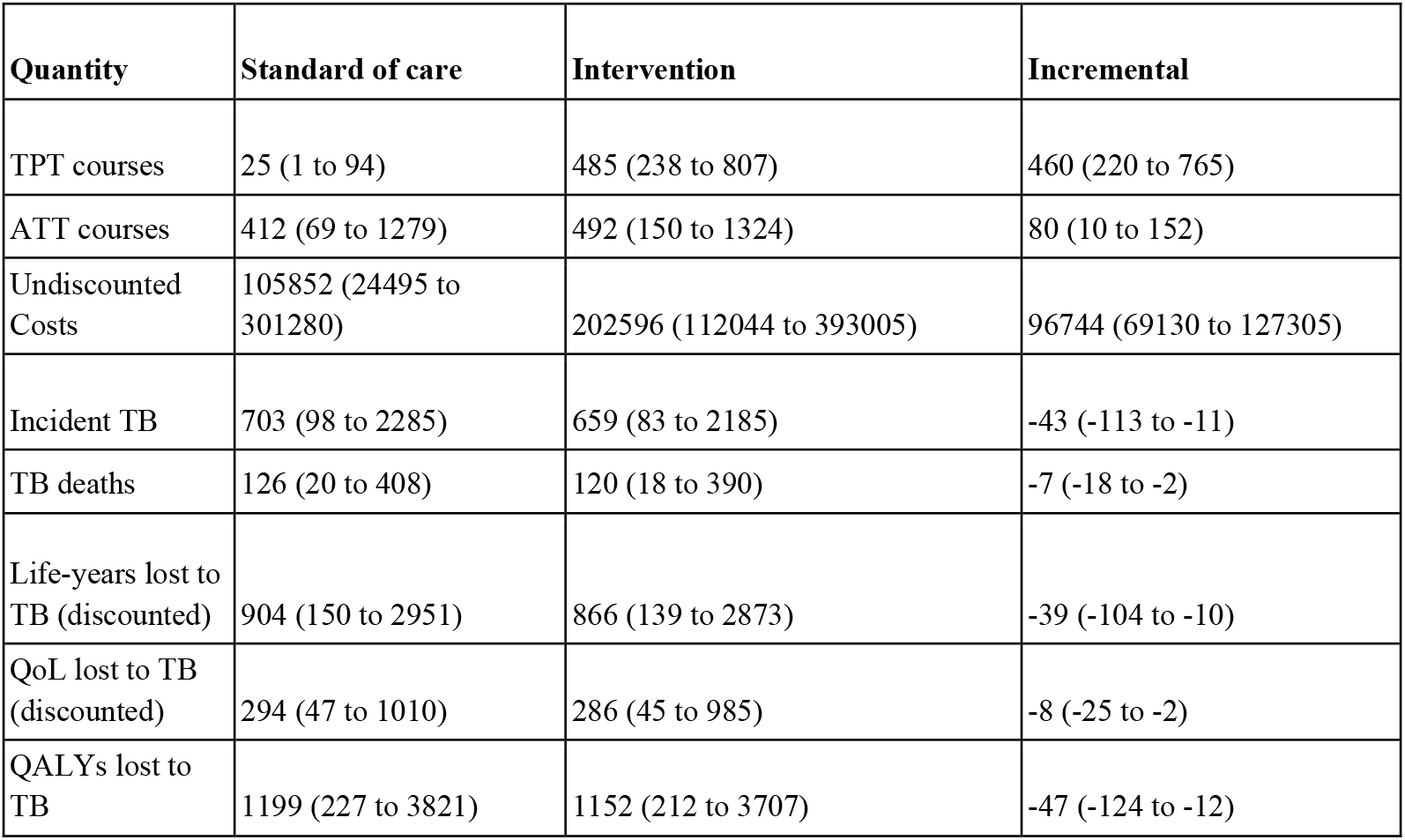
Resource use and health benefits per 10,000 people. TPT = tuberculosis preventive therapy; ATT = antituberculosis therapy; TB = tuberculosis; QoL = quality of life (due to tuberculosis morbidity but not mortality); QALYs = quality-adjusted life-years (including both life-years lost and morbidity). Parentheses denote 95% quantiles around mean.

| **Quantity** | **Standard of care** | **Intervention** | **Incremental** |
| --- | --- | --- | --- |
| TPT courses | 25 (1 to 94) | 485 (238 to 807) | 460 (220 to 765) |
| ATT courses | 412 (69 to 1279) | 492 (150 to 1324) | 80 (10 to 152) |
| Undiscounted Costs | 105852 (24495 to 301280) | 202596 (112044 to 393005) | 96744 (69130 to 127305) |
| Incident TB | 703 (98 to 2285) | 659 (83 to 2185) | -43 (-113 to -11) |
| TB deaths | 126 (20 to 408) | 120 (18 to 390) | -7 (-18 to -2) |
| Life-years lost to TB (discounted) | 904 (150 to 2951) | 866 (139 to 2873) | -39 (-104 to -10) |
| QoL lost to TB (discounted) | 294 (47 to 1010) | 286 (45 to 985) | -8 (-25 to -2) |
| QALYs lost to TB | 1199 (227 to 3821) | 1152 (212 to 3707) | -47 (-124 to -12) |

We projected the intervention to result in substantial increases in TPT use: an increase of 460 (95% uncertainty interval [UI]: 220 to 765) courses per 10,000 people eligible for screening (Table 2). The net effect on ATT courses was a smaller increase of 80 (95%UI: 10 to 152) courses per 10,000 people eligible for screening, despite a reduction in tuberculosis incidence of 43 (95%UI: 11 to 113) per population. These increases in treatment, together with the increased resources for screening and diagnosis resulted in a net increase in economic costs of £96,744 (95%UI: £69,130 to £127,305) per 10,000 people screened.

These efforts were projected to result in reduced mortality of 7 (95%UI: 2 to 18) per 10,000 people screened, and gains in HRQoL of 8 (95%UI: 2 to 25) per 10,000 people screened, which together implied gains of 47 (95%UI: 12 to 124) QALYs. Comparing the increased resources and health gains produced a base case ICER of £78,000 per QALY gained, in excess of the usual UK cost-effectiveness thresholds (Table 2). The sensitivity analysis assuming no LTFU yielded an ICER of £70,000/QALY gained; the sensitivity analysis assuming no escort costs in care cascades yielded an ICER of £54,000/QALY gained. The sensitivity analyses neglecting transmission and post-tuberculosis effects resulted in higher ICERs (see Appendix). The analysis variants restricting to tuberculosis notification rates in the range 30-100 per 100,000 resulted in higher ICERs: £168,000/QALY gained for the base case; £153,000/QALY gained for no LTFU; £118,000/QALY gained for no escort costs.

Exploring a range of target group size and tuberculosis infection prevalence for a zero-cost pre-screen, showed that higher target group tuberculosis infection prevalence and smaller target group size resulted in larger net benefit (Figure 3) Biobehavioral survey data suggested that tuberculosis infection prevalence was 20% in target group 1) and 8% in target group 2). These groups were estimated to comprise 6% and 20% of the screening group, respectively (Appendix). These values correspond to a positive net benefit for target group 1), which was cost saving (Figure 3). Targeting group 2), the net benefit was negative, and therefore not cost-effective at a threshold of £30,000/QALY gained (Figure 3).

**Figure 3:**
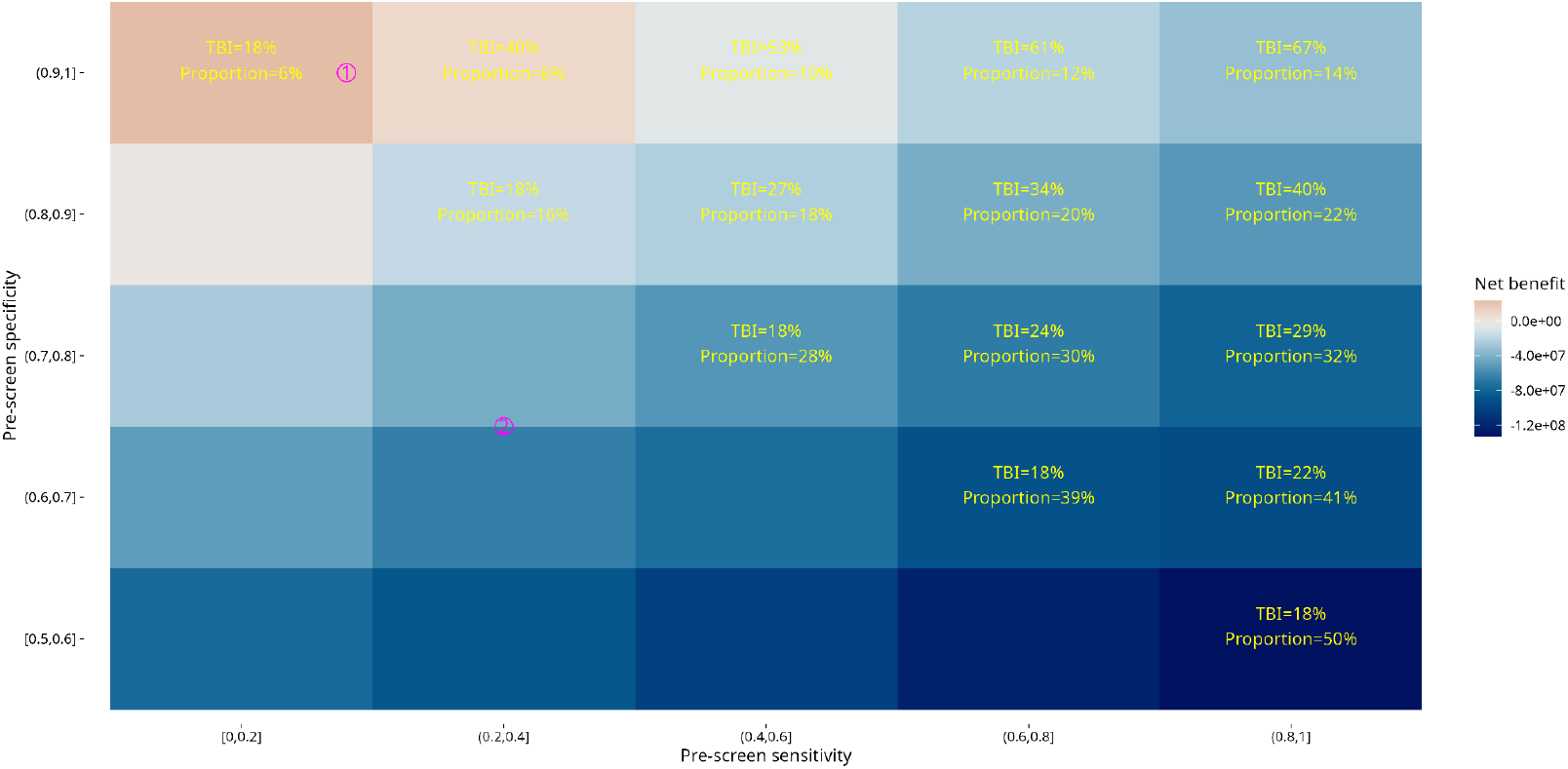
Net benefit for targeted screening approaches For each pre-screen sensitivity and specificity midpoint, yellow text shows the tuberculosis infection (TBI) prevalence and proportional size of the group screened-in for further assessment. Fill colour shows the mean expected net benefit for each tile, with red colours corresponding to positive net benefit (i.e. cost-effective at a threshold of £30,000 per quality-adjusted life-year gained); blue colours correspond to negative net benefit. Magenta numbers ①, ② locate the likely location of exemplar target groups defined in the text.

## Discussion

Our model-based cost-effectiveness analysis of tuberculosis infection screening and TPT on first reception into prisons in England found that non-targeted screening would not be cost-effective at typical UK cost-effectiveness thresholds. However, we found that targeting interventions using a zero-cost pre-screening step to select who is screened further has the potential to yield a cost-effective intervention. One specific example we considered, parametrized with survey data – namely applying the intervention to those born in medium or high tuberculosis incidence countries – resulted in an intervention that was not only cost-effective, but cost-saving due to tuberculosis management costs averted in prisons and the community.

The low cost-effectiveness of the intervention when applied without targeting stems from the relatively low prevalence of tuberculosis infection in English prisoners, the difficulty of achieving high completion of TPT in this population, and the high cost of prison-specific elements of the cascade of care. Recent survey data suggest that prisoners in England have IGRA positivity rates of 7%.^13^ While this prevalence is substantially higher than the general population, it remains below the estimated global average, and much below estimates for incarcerated individuals in high tuberculosis incidence settings. Using drop-offs in our modelled care cascade based on pilot data (Figure 2), we found that only 38% of those offered TPT ultimately received and completed a course. Finally, any care components that require prisoners to be securely escorted out of detention for treatment or assessment in hospitals are extremely costly due to costs associated with required prison staff escorts and bedwatch duties, and contribute to unit costs for assessment and treatment being higher than seen in the community.

Our analysis had limitations concerning the data available to parametrize flows through the prison system, the epidemiology of tuberculosis, and for parameterizing exploration of targeted screening. We did not have data on durations and flows between categories of UK prison, but instead made use of publicly available data and a Bayesian approach that dealt with the underdetermined nature of the problem. Doing this allowed a prison model that captured overall durations of detention, but would be less reliable for interventions targeted by sentence or prison type. We were not able to model the fraction of the inflow who had previously been incarcerated, nor were we able to consider targeted interventions focussing on those entering prison with a history of homelessness or substance misuse separately. There was limited data to inform tuberculosis transmission within prisons, and community incidence and transmission of tuberculosis associated with previously incarcerated individuals. However, our transmission model did take into account reductions of transmission within prison and subsequent community transmission was represented over a single generation. While a single generation of community transmission may miss some benefits, it is likely to capture most indirect effects due to declining tuberculosis rates and the use of discounting for costs and benefits. Furthermore, we explored many of these features with sensitivity analyses and found our conclusions qualitatively unchanged.

A more complete understanding of individuals’ overlapping social risk factors and patterns of engagement with health and social care over time would allow a better assessment of the interaction between public health policies aiming to engage at different entry points. This understanding would allow assessment of broader strategies for improving health and reducing inequalities for these high risk groups in a coordinated fashion.

Although there have been no previously published economic evaluations considering incarcerated individuals in the UK, cost-effectiveness analyses have been undertaken in other settings. In 2012, Winetsky et al.^18^ applied a transmission model to consider active case finding (ACF) strategies in the prisons in the former Soviet Union, with very high tuberculosis prevalence and extremely high rates of drug-resistance, and found that annual ACF rounds for tuberculosis disease were optimal. Since then, Kim et al^19^ considered digital X-ray for tuberculosis disease screening among people entering detention in South Africa, finding an ICER of 22,000 USD using a decision tree model. Smit et al^20^ considered screening for tuberculosis disease in Belgium finding ICERs of around 12,000 EUR per person identified with tuberculosis disease. The most comparable existing study to ours is Jo et al^21^, which undertook a cost-utility analysis of TPT in various target groups including prisoners for several US states using a transmission model. Jo et al. found ICERs ranging between 43,000-110,000 USD/QALY excluding New York, but did not consider prison-specific costs of care. A major strength of our work is a costing based on detailed prison-specific pathways of care, which account for the specific details of healthcare delivery in prisons.

Key challenges remain in designing effective and cost-efficient interventions in prison populations. Although pilot data exist, the proportion of people who will accept screening, and how this varies with risk is poorly known. Designing approaches which are acceptable and maximize participation is a priority. High rates of churn and short durations of detainment in remand mean investigations need to be rapid or have good referral linkages so as not to compromise completion rates. Costs associated with escorted trips out of prisons, which are also inefficient due to high cancellation rates, may be reduced by use of mobile x-ray kit and teams that can visit prisons and follow-up by telemedicine appointments. Finally, combining screening interventions across other conditions such as hepatitis C virus etc. may allow for economies of scope that bring down costs.

People in contact with the criminal justice system are an inclusion group identified for prioritization as part of NHS England’s Core20PLUS approach to reducing health inequalities. Moreover, people incarcerated in the UK have the right to an equivalent level of healthcare under the Mandela rules, the Council of Europe European Prison Rules, and the WHO framework on Health in Prisons.^22^ In order to achieve equivalence of care, the additional costs of surmounting the particular barriers and meeting the additional needs of those under the care of the state within the prisons system may require different willingness to pay than interventions for the general population.

Tuberculosis screening for those born in medium and high tuberculosis incidence countries entering prison is expected to be cost-saving as well as benefiting health. Further work should seek to design other targeted screening approaches that are also cost-effective. Cost-effectiveness could also be improved by approaches to screening and provision which increase acceptability and completion, and through delivery models that reduce prison-specific costs.

## Data Availability

All data produced are available online at https://github.com/nmafirakureva/PPDpathways

## Contributors

Designed and implemented analysis: NM, PJD, with input from all authors. First draft of article: NM, PJD. Data analysis NM, PJD, SW, CFF. Review of methods and results: RH, CFF, TF, AR, CE. Edited article and consented to submit: all authors. NM and PJD had full access to all the data in the study and had final responsibility for the decision to submit for publication.

## Data sharing

All code and data to reproduce this analysis are publicly available on GitHub at https://github.com/nmafirakureva/PPDpathways. The transmission model used in this analysis is available as an R package on GitHub at https://github.com/petedodd/ecrins.

## Acknowledgements

We would like to thank Lucy Thomas, Sharon Cox, Donna Foster, and Michelle Hodkinson for advice on data and input on analyses.

